# Calcitriol treatment is safe and increases frataxin levels in Friedreich Ataxia patients

**DOI:** 10.1101/2023.10.28.23297713

**Authors:** Berta Alemany Perna, Jordi Tamarit, Daniel López Domínguez, Anna Quiroga-Varela, Miguel Merchan Ruiz, Lluís Ramió i Torrentà, David Genís, Joaquim Ros

## Abstract

**Background:** Calcitriol, the active form of vitamin D (also known as 1,25-dihydroxycholecalciferol), improves the phenotype and increases frataxin levels in cell models of Friedreich Ataxia (FRDA).

**Methods:** Based on these results, we started a pilot clinical trial in which a dose of 0.25mcg/24h was administered to 15 FRDA patients for a year. Evaluations of neurological function changes (SARA scale, 9-HPT, 8-MWT, PATA test) and quality of life (Barthel Scale and SF-36 quality of life questionnaire) were performed. Frataxin amounts were measured in isolated platelets obtained from these FRDA patients, from heterozygous FRDA carriers (relatives of the FA patients) and from non-heterozygous sex and age matched controls.

**Results:** Although the patients did not experience any observable neurological improvement, there was a statistically significant increase in frataxin levels from initial values, 5,6 pg/μg, to 7,0 pg/μg after 12 months. Differences in frataxin levels referred to total protein amounts were observed amongst sex and age matched controls (18.11 pg/μg), relative controls (10.07 pg/μg), and FRDA patients (5.71 pg/μg). The treatment was well tolerated by most patients, and only some of them experienced minor adverse effects at the beginning of the trial.

**Conclusion:** Calcitriol dosage used (0.25mcg/24h) is safe for FRDA patients and it increases frataxin levels. We cannot rule out that higher doses administered longer could yield neurological benefits.

## INTRODUCTION

Friedreich ataxia (FRDA) is the most common autosomal recessive ataxia, caused by homozygous expansion of GAA trinucleotide repeat in FXN gene (*1, 2*) or, less frequently, by an heterozygous compound mutation of GAA repeat and a point mutation in the same gene (*3, 4*). Clinical manifestations are progressive limb and gait ataxia due to cerebellar atrophy, axonal polyneuropathy and dorsal cord syndrome. Additional features could include scoliosis, diabetes and hypertrophic cardiomyopathy, being the latter the main cause of death in FRDA patients (*5, 6*). The symptoms usually start in childhood or adolescence, but they can appear later (*7, 8*). The mutation results in low frataxin levels, being the main cause of the neurodegeneration. Although the exact function of frataxin is still a matter of debate, its deficiency has been associated with abnormal iron-sulfur cluster enzyme activities, mitochondrial iron accumulation and impaired mitochondrial function (*9–11*). Calcitriol, which is the vitamin D active form, is synthesized inside the mitochondria by means of cytochrome CYP27B1, a heme-containing hydroxylase acting on the precursor form, 25-OH-vitamin D3 (or calcidiol) and rendering 1,25-(OH)2-vitamin D3 (*12*). CYP27B1 function relies on the activities of ferredoxin 1 (FDX1), a Fe/S cluster-containing protein, which transfers electrons from a ferredoxin reductase (FDXR) to the cytochrome (*13*–*15*). Calcitriol maintains cellular redox balance and calcium signaling pathways as it contributes to regulate the expression of Nrf2 and Klotho (*16–18*). Besides these functions, calcitriol exerts, to cite some examples, protective effects against glutamate-induced excitotoxicity in hippocampal cells (*19*) and delays amyotrophic lateral sclerosis (ALS) progression by inducing axonal regeneration (*20*). Moreover, this compound has been approved for the treatment of renal chronic insufficiency, hypoparathyroidism and vitamin D-dependent or resistant rickets (*21*).

A recent study performed in primary cultures of frataxin-deficient dorsal root ganglia neurons, found that the amount of FDX1 protein was reduced by 50% of the control levels; interestingly, lymphoblastoid cell lines obtained from FRDA patients showed a 60% reduction in FDX1 levels compared to cells obtained from healthy donors (*22*). Of note, supplementing dorsal root ganglia neuronal cultures with 20nM calcitriol, restored altered parameters such as mitochondrial membrane potential, calcium homeostasis and improved neuronal survival. Moreover, calcitriol was also able to significantly increase frataxin amounts in both frataxin-deficient DRG neurons and lymphoblastoid cell lines (*22*). Based on the already well-known properties of calcitriol and the recent findings mentioned above, we performed a pilot clinical trial in which a low dose of calcitriol was administered to 15 FRDA patients for a year. In these patients we measured whether frataxin levels in platelets increased and the effect and safety of low doses of calcitriol on neurological function and quality of life.

## METHODS

### Study design

The study took place over 12 months. It was an open, without placebo clinical trial, and it was conducted in Hospital Josep Trueta/Hospital Santa Caterina in Girona, Spain, between September 2021 and September 2022.

The inclusion criteria were 1) being between 16 and 65 years of age, 2) having genetically confirmed FRDA (two pathological GAA triplet repeats in the FXN gene or one pathological GAA triplet repeat and one point mutation in the FXN gene) and 3) having retained the ability to walk with any kind of external aid (scoring SARA gate item ≤ 6). Women were asked to use an effective contraceptive method during the trial. The exclusion criteria include severe visual or auditory loss, cognitive decline, serious psychiatric illness or substance abuse, cardiac disease with heart failure or symptomatic valvular or coronary artery disease or being affected by any neurological or any other kind of disease that could interfere with the trial. The patients could not participate in other clinical trials or be under treatment with drugs that could raise calcium concentration levels (i.e., digoxin, thiazide diuretics, cholestyramine, corticoids, laxatives with magnesium, barbiturates and antiepileptic drugs). Women should not have a positive pregnancy test or be breastfeeding. Having hypercalcemia or elevated creatinine prior to the beginning of calcitriol treatment were also exclusion criteria. Developing hypercalcemia during the course of the trial meant withdrawal from the study. Another withdrawal criterion was being unable to follow the treatment indications and the appointments.

Every patient conducted 6 appointments to assess the neurological function and to control the hypercalcemia risk: at the start of the study (t0), at fifteen days (t0.5), and in the 4^th^ month (t4), 6^th^ month (t6), 8^th^ month (t8) and 12^th^ month (t12). The neurological scales used in the study were the SARA scale, the 9-Hole Peg test (9-HPT), the 8 Meters Walking Test (8-MWT) and the PATA velocity test. These tests were performed at the t0, t6 and t12 appointments. The patients’ quality of life and daily life activities were also examined with the Barthel scale and the SF-36 questionnaire at the t0 and t12 appointments.

An electrocardiogram and a blood analysis control were performed in all the appointments, except for the 6^th^ month appointment. Frataxin levels were tested at t0, t0.5, t4 and t12. Frataxin levels were also measured in FRDA carriers (either a heterozygous sibling or one of both parents) and age and sex matched controls with no known relatives affected by FRDA. The controls could not have any neurological disease or a condition related with calcium metabolism, and could not be under treatment to calcium or vitamin D.

The trial received the approval of the AEMPS (Spanish Agency for Medication and Healthcare Products) with the EudraCT number 2020-001092-32, and it also received the approval of the hospital Local Committee (CEIm Girona). The trial was registered in Clinicaltrial.gov (identifier NCT04801303)

### Primary and secondary endpoints

The main objective of the trial was to evaluate the effects of calcitriol on the neurological symptoms of patients with FRDA. The SARA scale, 9-Hole Peg test (9-HPT), 8-Meters Walking Test (8-MWT) and PATA velocity-test (PATA test) were used to assess changes in neurological functions.

There were three secondary objectives. The first was to assess the treatment safety and the hypercalcemia risk with low doses of calcitriol (0.25mcg of calcitriol every 24h), by performing basal and control electrocardiograms and blood tests with calcium and renal function. The second was to control the effects of calcitriol on daily life activities and the life quality of FRDA patients, which were assessed with the Barthel Index for Activities of Daily Living (Barthel scale) and the Quality-of-life test (SF-36 questionnaire). The third was to measure whether there was an increase of frataxin levels during the calcitriol treatment as it occurred in the cell cultures. To validate the frataxin levels measurement, a blood analysis was also done in two kinds of controls for every FRDA patient: a heterozygous FRDA control, and a healthy age- and sex-matched control.

### Frataxin levels measurements

Venous blood from donors (patients, carriers or controls) was drawn into two 8.5 mL ACD (Citric acid; Na-Citrate; Dextrose) Vacutainer tubes. Platelets were obtained using published procedures (23). Briefly, tubes were centrifuged at 200xg for 13 min at room temperature (with no brake applied). After the spin, two-thirds of the top layer (the platelet rich plasma or PRP) was transferred into a new tube using a wide orifice transfer pipette and centrifuged at 800 x g for 10 min (at room temperature with no brake applied). Supernatant was discarded and 1 ml of PBS containing 1ug/ml Prostaglandin E1 (Sigma-Aldrich P5515) was added to the pellet, without resuspending it. This preparation was centrifuged at 900 x g for 6 min, at room temperature with no brake applied. Supernatant was discarded and the pellet was resuspended in 250 ul of lysis buffer (Tris 0.1 mM pH 6.5, 3% SDS, EDTA 6.8 mM). This platelet lysate was transferred to a screw-capped 1.5 ml polypropylene tube. Tubes were shipped to the analytical laboratory in dry ice. Once there, they were heated at 95 ºC for five minutes, sonicated for five minutes in an ultrasonic bath at room temperature and centrifuged at 10000xg for five minutes to discard any insoluble material. Protein concentration in the supernatant was calculated on the basis of the absorbance at 280 and 260 nm (measured in an Implen NanoPhotometer N60). Samples were aliquoted and stored at -80 ºC until their analysis by western blot. For this analysis, 11ug of protein was mixed with 230 pg of pure recombinant His-tagged frataxin (Abcam ab95502, used as an internal standard), 3% beta-mercaptoethanol, and loaded onto 12.5% polyacrylamide SDS-PAGE gels. After electrophoresis, proteins were transferred to nitrocellulose membranes and frataxin was detected using FXN-specific (Abcam, ab219414, rabbit) and a CF™ 640R conjugate secondary antibodies (Sigma, SAB4600399). Images were acquired using a ChemiDoc MP system and analysed with ImageLab software (Bio-Rad). Frataxin was measured in samples obtained from healthy donors, carriers and patients at basal (before starting calcitriol treatment) and at 15 days, 4 months and 12 months of treatment.

### Statistical analysis

The planned number of subjects of this pilot trial was 20. It was not required a formal calculation of the sample size due to the kind of study design. In this regard, in the general rules to determine the appropriate sample size for a pilot study have been described in the published scientific literatura (24-27). It is recommended a recruitment of 24-40 patients for a pilot study with two intervention groups. For a pilot study with only one group, the calculation can be adapted with an adjustment factor of 0,5. Hence, in our study it was estimated that a minimum recruitment of 15-25 subjects would be needed, assuming a loss ratio of 20%.

Categorical variables were expressed as absolute and relative frequencies and continuous variables as means and standard deviations, or as medians and interquartile range if the distribution was not normal. The normality of the distribution of the variables involved was checked by the Shapiro-Wilk test.

To assess the effect of calcitriol treatment on neurological function and the evolution of frataxin levels, Student’s t-test for pairs (normal distribution) or the Wilcoxon test (non-normal distribution) were used. To compare frataxin levels amongst the different groups (healthy controls, heterozygous controls and patients) a one-way analysis of variance (ANOVA) or Kruskal-Wallis test was performed, based on the normality of the variables. We performed a preliminary analysis to test the hypotheses of normality and homoscedasticity. The Student’s t-test or the Mann-Whitney test were used for post-hoc pairwise comparisons. For multiple comparison a Bonferroni correction was performed. Regression line from the fold-change in frataxin expression was calculated by GraphPad Prism Software and the P value was calculated from an F test.

The analyses were performed using statistical software R version 4.2.1 with the “rstatix” and “compareGroups” packages. All *p*-values were 2-sided, and *p* less than 0.05 was considered statistically significant.

## RESULTS

The initial pool of participants were 20 patients (10 female; 10 male) aged from 16 to 55 years. Five patients could not complete the one-year treatment because of minor hypercalcemia and therefore were withdrawn from the study. The final sample of participants were 15 patients, who received a daily dose of Rocaltrol^©^ (calcitriol) 0.25mcg/24h for 53 weeks.

All participants met the inclusion criteria and signed the consent form to participate in the study.

### Clinical measures

The primary objective of this research project was to evaluate the neurological changes in FRDA patients treated with low doses of calcitriol. Neurological symptoms were assessed with SARA scale, 9-Hole Peg test (9-HPT), 8-Meters Walking Test (8-MWT) and PATA velocity-test (PATA test).

There were no significant changes in the SARA scale scores (Table 1). The total SARA values increased an average of 0,73 points in one year (t0 14.8, t12 15.53, *p* =0.147), indicating a neurological deterioration.

**Table 1.** SARA score evolution for the 15 patients that completed the one-year treatment with calcitriol. In the t0 column are expressed the total score and individual item score before the beginning of the treatment and in the t12 the total score and individual item score after one-year treatment. Scores are expressed with the mean and the (standard deviation) when the variable follows the normal distribution or with the median and the [interquartile range] when the variable does not follow a normal distribution.

| SARA score (n=15) | t0 | t12 | P value |
| --- | --- | --- | --- |
| SARA total score | 14.8 (±6.8) | 15.5 (±6.2) | .133 |
| SARA gait | 4 [2;4.5] | 5 [2;6] | .034 |
| SARA stance | 2 [2;3] | 2 [2;3] | .317 |
| SARA sitting | 1 [0.5;1] | 1 [1;1.5] | .025 |
| SARA speech disturbance | 1 [1;2] | 1 [1;2] | 1 |
| SARA finger-chase | 1.5 [1;1.75] | 1 [1;1.5] | .011 |
| SARA nose-finger test | 1 [0.25;1.25] | 1 [0.5;1] | 1 |
| SARA fast alternating hand movements | 1.5 (±0.8) | 1.267 (±1.0.) | .169 |
| SARA heel-shin | 2.5 [2;3] | 2.5 [2;3] | .305 |

Similarly, there were no observed significant changes in the 9-HPT (t0 53,94 seconds, t12 56.90 seconds, *p* = .120) and the PATA test (t0 21,25 seconds, t12 22.25 seconds, *p* = 0.477). The patients experienced a significant worsening of the 8-MWT (t0 22,71 seconds, t12 36.51 seconds, *p* = 0.0067) (Table 2) that falls in line with the observed variation in the “Gait item” of the SARA scale (t0 4, t12 5, *p* = 0.0337). One patient was not able to complete the 8-MWT.

**Table 2.** 9-HPT total scores and scores for DM (dominant hand) and NDH (non-dominant hand), 8-MWT total scores and scores for trial 1 (8 meters going-way) and trial 2 (8 meters returning-way), and PATA test score changes for the 15 patients that completed the one-year treatment with calcitriol. In the t0 column are expressed the score just before starting the treatment and in the t12 column the score after one-year treatment. Scores are expressed with the median and the [interquartile range] since the variables do not follow a normal distribution.

| **9-HPT score (n=15)** | **t0** | **t12** | **P value** |
| --- | --- | --- | --- |
| 9-HPT total score (seconds) | 53.94<br>[51.475;64.255] | 56.90<br>[51;73.185] | .120 |
| 9-HPT DM (seconds) | 50.98<br>[42.52;57.945] | 54.45<br>[46.455;62.66] | .761 |
| 9-HPT NDM (seconds) | 60.27<br>[53.085;70.67] | 61.52<br>[54.045;86.355] | .073 |
| **8-MWT score (n=14)** | **t0** | **t12** | **P value** |
| 8-MWT total score (seconds) | 22.71<br>[15.685;36.335] | 36.51<br>[18.925;57.785] | .007 |
| 8-MWT trial 1 (seconds) | 12.58<br>[7.7125;17.8575] | 15.41<br>[9.375;25.7475] | .008 |
| 8-MWT trial 2 (seconds) | 14.44<br>[9.2725;21.3225] | 21.69<br>[9.05;32.0375] | .091 |
| **PATA test score (n=15)** | **t0** | **t12** | **P value** |
| PATA test total score<br>(number of PATA syllables) | 21.25<br>[18.75;23.25] | 22.25<br>[19.875;25] | .477 |

The score in the Barthel scale remained steady (t0 and t12 85%, *p* = 0.144) and the patients did not experience any significant variation in the quality of life questionnaire (t0 55.99%, t12 59.336%, *p* = 0.89) (Table 3). The only exception was in the overall health perception in the SF-36 questionnaire, in which patients were asked to rate their health on a 5-point scale at the time the questionnaire was delivered, compared to one year prior. At the end of the study, patients perceived their health condition had significantly improved by one point, from ‘Somewhat worst now than one year ago at the beginning of the study to ‘About the same’ at the end of the clinical trial (t0 25%, t12 50%, *p* = 0.019).

**Table 3.**
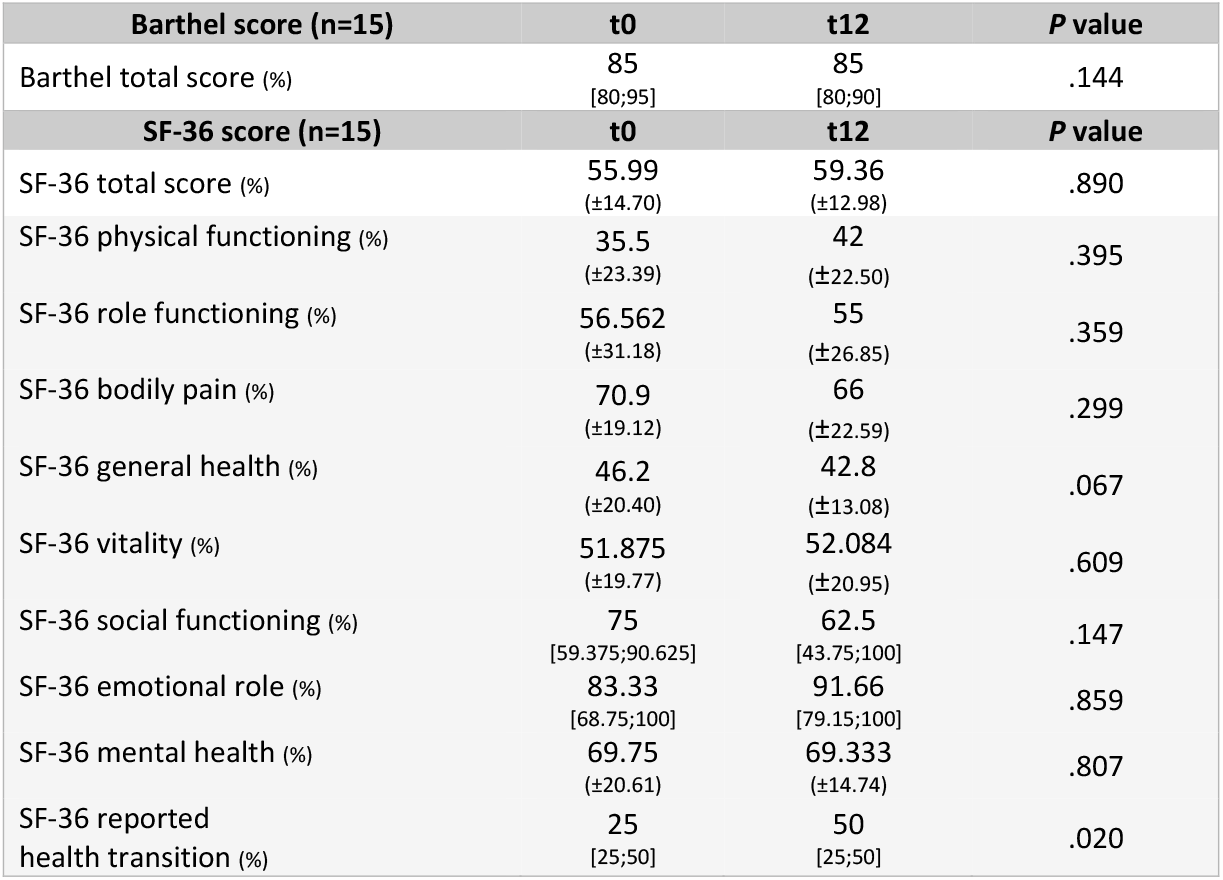
Barthel scale total score and SF-36 health-survey total score and individual item scores variations for the 15 patients that completed the one-year treatment with calcitriol. In the t0 column are expressed the scores before the beginning of the treatment and in the t12 column the scores after one-year treatment. Scores are expressed with the mean and the (standard deviation) when the variable follows the normal distribution or with the median and the [interquartile range] when the variable does not follow a normal distribution.

| **Barthel score (n=15)** | **t0** | **t12** | **P value** |
| --- | --- | --- | --- |
| Barthel total score (%) | 85<br>[80;95] | 85<br>[80;90] | .144 |
| **SF-36 score (n=15)** | **t0** | **t12** | **P value** |
| SF-36 total score (%) | 55.99<br>(±14.70) | 59.36<br>(±12.98) | .890 |
| SF-36 physical functioning (%) | 35.5<br>(±23.39) | 42<br>(±22.50) | .395 |
| SF-36 role functioning (%) | 56.562<br>(±31.18) | 55<br>(±26.85) | .359 |
| SF-36 bodily pain (%) | 70.9<br>(±19.12) | 66<br>(±22.59) | .299 |
| SF-36 general health (%) | 46.2<br>(±20.40) | 42.8<br>(±13.08) | .067 |
| SF-36 vitality (%) | 51.875<br>(±19.77) | 52.084<br>(±20.95) | .609 |
| SF-36 social functioning (%) | 75<br>[59.375;90.625] | 62.5<br>[43.75;100] | .147 |
| SF-36 emotional role (%) | 83.33<br>[68.75;100] | 91.66<br>[79.15;100] | .859 |
| SF-36 mental health (%) | 69.75<br>(±20.61) | 69.333<br>(±14.74) | .807 |
| SF-36 reported health transition (%) | 25<br>[25;50] | 50<br>[25;50] | .020 |

### Side effects

Side effects were assessed through patient interviews, electrocardiograms and blood control analyses. Results showed that neither moderate nor severe side effects occurred during the treatment.

Some patients complained about mild side effects, mainly during the first 15 days of treatment. Four patients had minor side effects that were solved in a few days (one had a headache, one had nausea, one had constipation and one had polaquiuria). At the 4^th^ month appointment only one patient reported a mild headache, which lasted for an hour after taking the drug, but this side effect lasted only one month. There were no more calcitriol side effects during the rest of the trial.

As mentioned above, five patients were withdrawn from the trial due to mild hypercalcemia. None of these patients reported side effects. The hypercalcemia levels were always below 10.3 mg/dL (normal calcium levels 8.6-10.1 mg/dL), which returned to normal levels a week after stopping the treatment.

### Frataxin quantitation

Frataxin levels were measured in platelets obtained from patients, carriers, and healthy donors. Platelets had previously been used to measure frataxin expression in FRDA patients, as they are easily accessible, contain active mitochondria and present a low half-life (*23-25*). Analysis was performed by western blot, using a recombinant His-tagged frataxin as an internal standard. In order to validate this method, frataxin was measured in platelets obtained from 16 patients, carriers, and healthy donors. As expected, frataxin content was significantly lower (p<.001) in patients (5.71 pg/μg) than in carriers (10.07 pg/μg) or healthy donors (18.11 pg/μg) (figure 1A), validating the accuracy of the assay.

**Figure 1.**
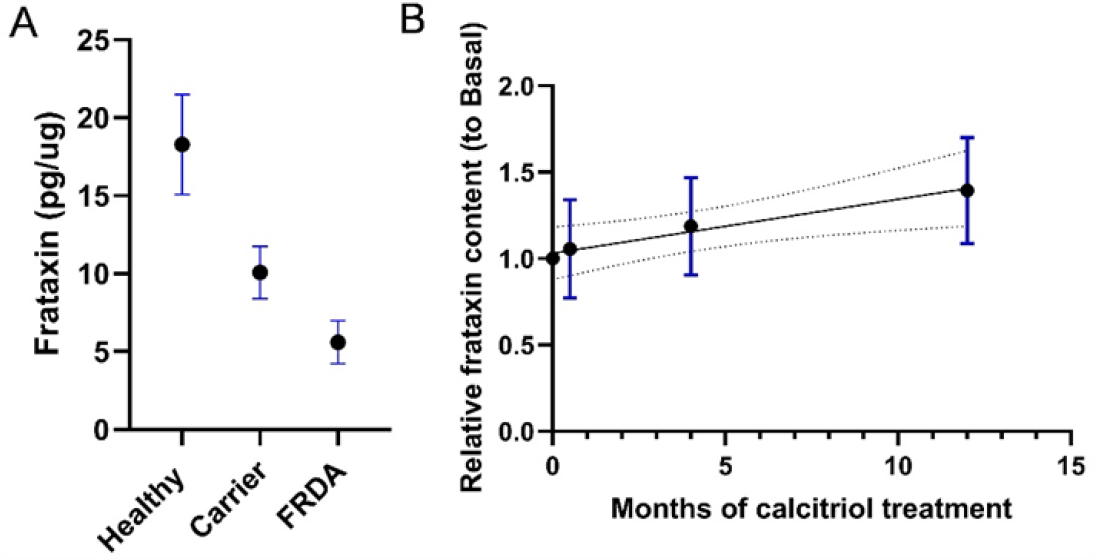
Calcitriol treatment increases frataxin protein content in FRDA patients platelets. A. Frataxin levels at baseline of 16 healthy, carrier and FA individuals. Mean +/- 95% confidence intervals are shown. B. Fold-change in frataxin expression relative to baseline observed over 12 months calcitriol treatment in 15 FRDA patients completing the trial. Slope from the regression line is significantly non-zero (p value 0,0112). Dashed lines indicate the 95% confidence bands of the regression line.

Results showed that frataxin levels increased gradually in the 15 patients under calcitriol treatment (figure 1B). There were significant differences (p = .007) in the Frataxin levels in the 15 patients that took calcitriol for a year, from 5,54 pg/ug (SD ±2.61) at the beginning of the treatment to 6.98 pg/ug at the end of the study (SD ± 3.21).

## DISCUSSION

### Effects of calcitriol in the neurological symptoms, daily life activities and patients quality of life

The calcitriol treatment did not improve patients neurological function, showing an increase of 0.73 points in the SARA score after one-year of treatment. The changes observed in this cohort of patients cannot be compared against a FRDA natural history, as there have been no studies measuring changes in the SARA scale in the timespan of one year. There have been, however, two studies carried out by The European Friedreich’s Ataxia Consortium for Translational Studies (EFACTS) which have assessed the rate of disease progression in FRDA patients after 2 and 4 years (*30, 31*.). They observed an average SARA scale progression rate of 0.77/year for 2-year study and 0.82/year for 4-year study. Since the present study only assessed patients over a year, it was not possible to calculate an annual progression rate, and thus comparison with the aforementioned studies should be made with care. That being said, the SARA scale scores in the present study showed that the patients progression (0.73/year) was similar to the progression rate calculated for the 2 years (0.77/year) and 4 years (0.82/year) studies. It is possible that the progression observed in the FRDA patients treated with calcitriol is similar to what would have been observed if the patients had not received the calcitriol treatment, suggesting that the treatment did not cause an exacerbation of the neurological deterioration in our FRDA patients.

There were neither changes in the daily life activities nor general perception of quality of life. The only exception was the patient perceived health condition, which improved after the one-year treatment. A reason for this change could be that patients were doing a more frequent follow-up, which could have helped them improve their health perception.

### Safety and hypercalcemia risk with low doses of Calcitriol

Results confirm that calcitriol at low dose is a safe drug for FRDA patients. Only minor side effects were observed (such as headache, nausea, constipation and polaquiuria), which resolved in few days or weeks. The only moderate side effect observed during the calcitriol treatment was low-level asymptomatic hypercalcemia in 25% of the patients, and levels returned to normality a week after patients stopped taking the drug. Thus, the minimal and manageable side effects, along with the reversible nature of hypercalcemia, highlight the drug’s safety and tolerability.

### Frataxin levels changes during the calcitriol treatment

One of the main findings of this work is that the levels of frataxin increase significantly during the calcitriol treatment, which suggest that calcitriol can promote the expression of frataxin in FRDA patients and may improve mitochondrial function. However, the treatment with calcitriol at 0.25mcg/24h did not cause significant neurological improvement. This falls in line with results from prior studies with other treatments in FRDA patients, which have also shown an increase in the frataxin levels (*28, 32*) but have not found any change in the neurological function (*33, 34*).

Few studies that have assessed the frataxin levels in FRDA patients. These studies have measured frataxin in fibroblasts, lymphocytes, muscle biopsies (35), buccal cells, peripheral blood (*36, 37*) and platelets (29, 38). These studies have shown that frataxin levels in peripheral tissues ranges from 30% to 40% of control levels in FRDA patients and from 60 to 80% of control levels in FRDA carriers (*39-42*). As we have observed similar differences between the three groups, we can conclude that the results obtained are accurate.

## CONCLUSIONS

Calcitriol, with a minimum dose of 0.25mcg/24h, increases frataxin levels in FRDA patients and causes minimum side effects, although the lack of significant neurological improvement highlights the intricate nature of FRDA progression. Potential neurological benefits with longer treatments and/or higher doses of the compound (for instance, 0,50 mcg daily which still is a normal dose) can be considered for future interventions.

### LIMITATIONS

The study has a few limitations that should be acknowledged. First, the sample size was small. Second, the clinical trial was an open trial without placebo. The decision to have an open trial without placebo was precisely due to the fact that obtaining a larger sample of FRDA patients was challenging. Similarly, the withdrawal of 5 patients during the clinical trial because of minimal hypercalcemia further reduced the sample size. Third, the patient sample was heterogeneous. Two patients had late onset FRDA and two patients had compound heterozygous mutations (one with a GAA repeat and a punctual mutation, and another with a GAA repeat and one deletion). The GAA length was not available from the genetic studies reported by the patients, and hence it was not possible to compare the frataxin levels with the genetic status of every FRDA patient. Further research with a larger sample and a longer follow up period could overcome these limitations.

## Data Availability

All data produced in the present study are available upon reasonable request to the authors

## FUNDING AND ACKNOWLEDGEMENTS

This research was funded by FEDAES (Federación de Ataxias de España) and by Ministerio de Ciencia e Innovación (Spain), grant number PID2020-118296RB-I00.

We thank Roser Pané for technical assistance. We acknowledge Juan Carlos Baiges for support and helpful discussions of the manuscript and Geòrgia Pujadas-Jorbà for her generous help with the manuscript translation.

Above all, we truly acknowledge the patients and their families for their participation and effort.

